# The genetic dissection of fetal haemoglobin persistence in sickle cell disease in Nigeria

**DOI:** 10.1101/2023.05.16.23289851

**Authors:** Oyesola O. Ojewunmi, Titilope A. Adeyemo, Ajoke I. Oyetunji, Bassey Inyang, Afolashade Akinrindoye, Baraka S. Mkumbe, Kate Gardner, Helen Rooks, John Brewin, Hamel Patel, Sang-Hyuck Lee, Raymond Chung, Sara Rashkin, Guolian Kang, Reuben Chianumba, Raphael Sangeda, Liberata Mwita, Hezekiah Isa, Uche-Nnebe Agumadu, Rosemary Ekong, Jamilu A. Faruk, Bello Y. Jamoh, Niyi M. Adebiyi, Ismail A. Umar, Abdulaziz Hassan, Christopher Grace, Anuj Goel, Baba P.D. Inusa, Mario Falchi, Siana Nkya, Julie Makani, Hafsat R. Ahmad, Obiageli Nnodu, John Strouboulis, Stephan Menzel

## Abstract

**Background:** The clinical severity of sickle cell disease (SCD) is strongly influenced by the level of fetal haemoglobin (HbF) persistent in each patient. Three major HbF loci (*BCL11A*, *HBS1L-MYB*, and *Xmn1-HBG2*) have been reported, but a considerable hidden heritability remains.

**Aim:** Building on the power of a large and genetically diverse patient pool present in Nigeria, we conducted a genome-wide association study for HbF levels in patients from three regions of the country with a diverse ethnic make-up.

**Methods:** We analysed genome-wide trait association in 1006 Nigerian patients with SCD (HbSS/HbSβ^0^), followed by a replication and meta-analysis exercise in four independent SCD cohorts (3,582 patients). To dissect association signals at the major loci, we performed stepwise conditional analysis, haplotype association analysis and included public functional annotation data (fGCTA).

**Results:** Association signals were detected for *BCL11A* (lead SNP rs6706648, β=- 0.39, *P*=4.96 x 10^-34^) and *HBS1L-MYB* (lead SNP rs61028892, β=0.73, *P*=1.18 x 10^-9^), whereas the variant allele for *Xmn1-HBG2* was found to be very rare. Genetically dissecting the two major loci, we defined trait-boosting haplotypes containing suspected or so-far unidentified causal variants. At *BCL11A*, one such haplotype (trait increase *P* < 0.0001) contains the putative functional variant rs1427407-‘T’ and a second haplotype (*P* < 0.0001) is tagged by the rs7565301-‘A’ allele, with no obvious candidate causal variant. At *HBS1L- MYB*, one haplotype (trait increase *P* < 0.0001) contains the likely functional rs66650371 (Δ3-bp), and a second (*P* < 0.0001) is tagged by the ‘C’ allele of rs6102889. Together, variants at *BCL11A* and *HBS1L-MYB* SNPs explained 24.1% of the trait variance.

We detected three novel association signals: *SLC28A3* on chromosome 9 (rs115555854: β=- 0.73, *P*=2.52 x 10^-8^), *TICRR* on chromosome 15 (rs140496989: β=-0.43, *P*=3.34 x 10^-8^), and *PIEZO2* on chromosome 18 (rs58817161: β= −0.63, *P*= 8.04 x 10^-8^). These appeared to be restricted to the Nigerian patient cohort and were not confirmed in the replication cohorts.

**Conclusions:** Studying a diverse cohort of Nigerian patients with sickle cell disease, we genetically dissected the known fetal-haemoglobin loci *BCL11A* and *HBS1L-MYB* and detected putative new trait-associated regions.

## Introduction

Sickle cell disease (SCD) is an inherited blood disorder that affects approximately 300,000 newborns globally every year. Half of these are born in Nigeria alone, where SCD is a major public health problem and contributes significantly to childhood morbidity and mortality as fifty percent of children do not live beyond their 5^th^ birthday [1].

Polymerisation of haemoglobin S (HbS) under hypoxic conditions is central to the pathophysiologic events in SCD, giving rise to many debilitating complications including vaso-occlusive crises, stroke, and leg ulcers.

A wide variability in severity and clinical presentation is driven partially by additional genetic factors, such as the genotypic state at the site of mutation (homozygous: Hb SS; compound heterozygous: Hb SC, HbS beta thalassaemia), and by genetic disease modifiers at the beta globin gene locus or elsewhere in the genome. To discover the underlying genes and genetic variants is highly desirable; however, the immense complexity of clinical presentations and pathological pathways involved has been a major obstacle. A major breakthrough has been the discovery that a substantial persistence of fetal haemoglobin (HbF, α_2_γ_2_) in certain patients can ameliorate the disease severity and improve survival [2]. This led to intense efforts to find ways to therapeutically induce HbF expression.

Some HbF persists naturally in adults, to variable extent, forming a quantitative trait with up to 89% heritability in the general population and 60-70% in SCD patients [3, 4]. Since it has been challenging to assemble sufficiently large and homogeneous SCD study cohorts, the first HbF loci, and thus the first SCD modifier genes, outside the beta globin gene cluster, HBS1 like Translational GTPase-*MYB* Proto-Oncogene (*HBS1L-MYB)* and B-cell lymphoma/leukaemia 11A (*BCL11A)* were discovered through studies in an extended family [5] and in large non-anaemic population samples [6–8]. They were subsequently replicated in SCD patient cohorts [9]. Together, the three major HbF loci, the β-globin gene cluster on chromosome 11 (*Xmn1-HBG2* - a gamma globin promoter polymorphism), *BCL11A* on chromosome 2, and *HBS1L-MYB* on chromosome 6, account for about 50% of HbF variability in healthy persons and 20-30% in individuals with SCD in many populations [7, 9–13]. Therefore, half of the overall trait heritability is still unexplained, probably due to a lack of statistical power to detect large numbers of rare or weak-effect variants. To overcome this, to discover novel HbF or SCD severity loci and to better dissect the known ones, statistical power can be enhanced by assembling larger patient cohorts, by combining existing studies, or by ‘genetic loading’, i.e., recruiting subjects with extreme trait expression or marked family history. Nigeria, with its large number of patients, provides unique opportunities in this respect. The enhanced haplotype resolution seen in African populations is expected to support fine-mapping and the exceptional ethnic diversity of Nigeria will allow the evaluation of genetic variants that are rare elsewhere. Here, we present a first genome-wide study of fetal haemoglobin in patients with SCD in Nigeria, searching for new loci and performing fine-mapping to identify candidate functional variants and gain further insight into HbF regulation and heritability in persons with SCD.

## Materials and methods

### Patients

One thousand one hundred and fifty-eight individuals with sickle cell disease (HbSS/HbSβ^0^) recruited from four sites in Nigeria: Lagos University Teaching Hospital, Lagos; Sickle Cell Foundation Nigeria, Lagos; University of Abuja Teaching Hospital, Abuja; and Ahmadu Bello University Teaching Hospital, Zaria (Supplementary table 1) were genotyped. Patients were excluded from this study if they were on hydroxyurea, younger than 5 years old, had blood transfusion in the preceding 3 months, or had HbSC/HbSβ^+^. A uniform study proforma was completed for data collection across the project sites.

Five millilitres venous blood sample was collected into an EDTA bottle and processed immediately for complete blood count and haemoglobin profiling. The buffy coat was stored at −20^0^C for subsequent DNA extraction. Fetal haemoglobin levels were determined using high performance liquid chromatography (Bio-Rad D-10; Bio-Rad Laboratories, Hercules, CA, USA). FlexiGene DNA extraction kit (Qiagen GmbH, Hilden, Germany) was used for DNA extraction from buffy coat following the manufacturer’s protocols.

### Genotyping and quality control

Genotyping was performed using the Infinium™ H3Africa Consortium Array containing ∼2.3 million markers. One thousand one hundred and fifty-eight samples were genotyped and processed in Illumina’s Genome Studio software (version 2.05) for variant calling following the COPILOT raw Illumina genotyping quality control (QC) protocols detailed in [14]. Seventy-seven samples with genotyping call rate less than 90% were excluded during the Illumina Genome Studio QC and no sample was excluded further due to sample quality as the genotyping call rate was 99.99%. Individual-level QC was carried out to exclude samples with sex discrepancies compared with X-chromosome derived sex, heterozygosity outliers (heterozygosity ± 3 SD from mean), and genetically identical individuals (identity by descent, pi-hat ∼ 1.0) (Supplementary table 2), retaining 1006 individuals for imputation and downstream analysis. Per-marker QC excluded SNPs with call rate less than 97%, minor allele frequency < 1%, SNPs that deviated from Hardy-Weinberg equilibrium (*P* < 10^-8^), leaving 1,925,391 autosomal SNPs and X-chromosome. The overall genotyping rate was 99.994%. Quality control was carried out using PLINK v1.90 (www.cog-genomics.org/plink/1.9/).

### Imputation

Filtered genotyped data were used for imputation in TOPMed Imputation Server for pre-phasing with Eagle version 2.4 and imputation using Minimac4 (https://imputation.biodatacatalyst.nhlbi.nih.gov/#!pages/home). During post-imputation quality control, SNPs with INFO score < 0.4 and non-biallelic SNPs were excluded, retaining 14,816,370 SNPs for data analysis.

### Replication data set and meta-analysis

To evaluate our putative novel loci and to explore the additional potential gained from assembling a larger dataset through international collaboration, we involved summary-statistical datasets from UK SCD patients studied by us (Gardner et al. 2018) and from HbF GWAS studies in SCD patients performed by researchers in Tanzania [15] and the US [16]. Details of the patients’ recruitment, genotyping/sequencing technologies and platforms, data quality control, imputation, and association analyses for the Tanzanian and African-American data sets have been fully described [15, 16].

The UK SCD GWAS dataset is described separately [17], *manuscript in preparation*). It involves 626 HbSS/Sβ^0^ patients and 179 HbSC patients of West African and African Caribbean descent, with sickle mutation genotype included as a covariate during the association analysis. The Tanzanian Muhimbili Sickle Cell Cohort GWAS dataset includes 1,187 sickle cell disease patients and the St Jude Children’s Research Hospital (Memphis, TN, US) GWAS dataset includes 584 African-American SCD patients. The UKSCD and Tanzania SCD data were analysed using GCTA software with ln [HbF%] as the trait, with correction for age and sex while the African-American data were analysed with EMMAX software, using square root–transformed HbF% data as the trait [18]. Due to the different transformations of %HbF, sample size weighted meta-analysis (Stouffer’s method in METAL software) [19] with genomic control correction was performed taken in the SCD patients in the UK, Tanzania, and African-American cohorts. To identify new association signals, a second meta-analysis was performed to include the Nigerian cohort.

### Fine-mapping

The functional genome wide complex trait analysis (fGCTA) pipeline runs GCTA-COJO on scaled summary statistics based on the presence or absence of relevant annotations. The annotations used in this analysis are: K562 regulatory elements derived using chromHMM algorithm from Roadmap Epigenome[20, 21], HUDEP-2 obtained from peak calls from Chip-Atlas[22, 23] and GATA1 ChIP-seq on human peripheral derived blood-erythroblast [24]. By use of Bayesian methodology, variants that overlap with the annotation have a decreased standard error and vice versa. This pipeline was implemented in C (software is available here: https://github.com/cgrace1978) and was converted into a prior probability which is combined with the p-value of the variant to generate a posterior probability.

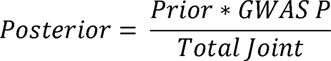

The posterior is then used to generate a new standard error (while keeping the original variant betas). These scaled summary statistics used GCTA-COJO module that performs a multi-SNP conditional and joint analysis of the summary data [25, 26]. The output of GCTA-COJO is independent variants in the summary statistics, which have a joint p-value below 1×10^-5^.

### Association analysis

To analyse data from a study population that is very diverse in terms of genetic/ethnic background, geographic origin, and age (median age of 15, range 5 - 60 years), we performed linear-mixed effects regression as implemented by GCTA (Genome-wide Complex Trait Analysis software, version 1.91.7) [25] to control for population stratification by incorporating genetic relationship matrix. This was done using the leave one chromosome out approach with age, sex, and recruitment sites as covariates. Natural logarithm of HbF was used to perform association testing. Manhattan and Q-Q plots were generated in R v.4.2.2 software using the qqman package [27]. *P*-value < 5.0 x 10^-8^ was regarded as being genome-wide significant.

### Conditional analysis and locus association plots

Conditional analyses were carried out with conditional and joint analysis in GCTA (GCTA-COJO) using summary statistics [26]. Regional association plots were generated in locus zoom [28].

### Haplotype association analysis

Haplotypes were constructed with Haploview 4.2 [29] using phased genotypes from TOPMed Imputation Server. Haplotype effects were estimated using ANOVA in R with adjustment for sex, age, and recruitment sites. Tukey’s post-hoc test was applied for multiple comparisons of means.

Data handling and statistical analyses were performed on the high-performance computing platform CREATE at King’s College London [30].

## Results

### Genome-wide association analysis in the Nigerian cohort

We conducted a genome-wide association analysis for 475 male and 531 female patients with sickle cell disease (HbSS/HbSβ^0^) recruited in Southwest, North-Central, and North-West Nigeria. The median HbF of the patients was 6.69 % (range: 0.8 – 32%) (Supplementary Table 1) and the population structure shows the diversity of the patients recruited for this study (Supplementary Figure 1).

We detected two of the three major HbF modifier loci (Figure 1, Table 1), *BCL11A* (lead SNP rs6706648, β = −0.39, *P* = 4.96 x 10^-34^) and *HBS1L-MYB* (lead SNP rs61028892, β = 0.73, *P* = 1.18 x 10^-9^). The third, *XmnI-HBG2* (rs7482144), was not included in the association analysis with our trait measure since this critical variant at the beta-globin gene cluster was rare (MAF = 0.007), consistent with a previous finding [31]. A list of genome-wide significant SNPs reported to be associated with HbF in previous studies and this current study is shown in Supplementary Table 3.

**Figure 1.**
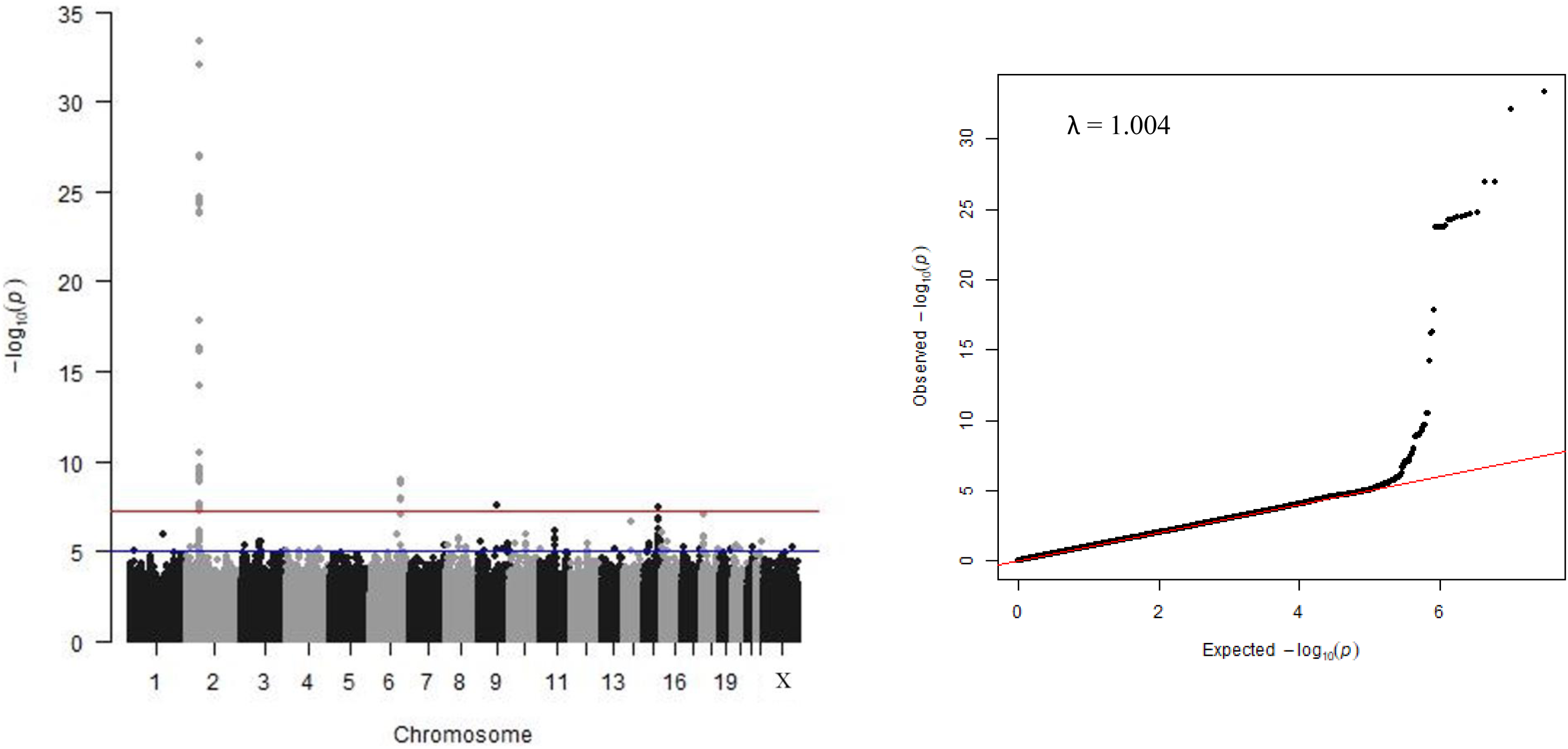
(a) **Manhattan plot of −log_10_(P-values)**. The red line indicates the threshold for genome-wide significant (*P* < 5 x 10^-8^), the blue line for suggestive association (*P* < 1 x 10^-6^). (b) **Quantile-Quantile plot of −log10(P-values) for all SNPs tested**. The red line indicates the null hypothesis that observed P-values equal expected P-values. Since there is no deviation from the null hypothesis line except at the tail end, population structure is implied to be effectively controlled. The low λ_GC_ value (lambda genomic control inflation factor) of 1.004 indicates that population stratification has been adjusted for effectively by the model and relationship matrix.

**Table 1.** List of genome-wide loci in Nigeria cohort and the replication cohorts.

| Chr | Locus | SNP | Position | EA/OA | Nigeria cohort = 1006 |  |  | Replication meta-analysis |  |  | Meta-analysis of Nigerian and replication cohorts |  |  |
| --- | --- | --- | --- | --- | --- | --- | --- | --- | --- | --- | --- | --- | --- |
| | | | | | EAF | $\beta$ | p-value | EAF | N | p-value | EAF | N | p-value |
| 2 | *BCL11A* | rs6706648 | 60494905 | T/C | 0.41 | -0.39 | $4.96 \times 10^{-34}$ | 0.42 | 2533 | $2.47 \times 10^{-60}$ | 0.42 | 3539 | $7.08 \times 10^{-92}$ |
| 6 | *HBS1L-MYB* | rs61028892 | 135097526 | C/G | 0.02 | 0.73 | $1.18 \times 10^{-9}$ | 0.02 | 2509 | $4.92 \times 10^{-17}$ | 0.02 | 3510 | $5.03 \times 10^{-25}$ |
| 9 | *SLC28A3* | rs115555854* | 84339933 | A/C | 0.01 | -0.73 | $2.52 \times 10^{-8}$ | - | - | - | 0.01 | 1003 | $2.73 \times 10^{-8}$ |
| 15 | *TICRR* | rs140496989 | 89619296 | A/G | 0.05 | -0.43 | $3.34 \times 10^{-8}$ | 0.04 | 2562 | 0.2216 | 0.04 | 3566 | 0.06 |
| 18 | *PIEZO2* | rs58817161 | 10818373 | C/T | 0.02 | -0.63 | $8.04 \times 10^{-8}$ | 0.02 | 2511 | 0.7841 | 0.02 | 3517 | 0.00197 |

Chr: Chromosome; SNP: reference ID for the Single Nucleotide Polymorphism; EA: Effect allele; OA: Other allele; EAF: Effect allele frequency; $\beta$ : allelic effect size; N = sample size. The replication meta-analysis includes SCD patients' datasets of UK, Tanzania, and African-American cohorts. Where the allelic effect size ( $\beta$ ) is positive, the HbF-boosting allele is the alternative allele; where it is negative, HbF is increased by the reference allele. Chromosomal positions are in hg38. \*MAF for rs115555854 was less than 1% in the UK, Tanzania, and African-American cohorts.

In addition, we detected two novel association signals at a genome-wide significance level: *SLC28A3* on chromosome 9 (*rs115555854*: β = −0.73, *P* = 2.52 x 10^-8^) and *TICRR* on chromosome 15 (*rs140496989*: β = −0.43, *P* = 3.34 x 10^-8^). A third locus, *PIEZO2* on chromosome 18, showed association at 5% false discovery rate (p-value < 1.37 x 10^-7^) (*rs58817161*: β = −0.63, *P* = 8.04 x 10^-8^) (Table 1).

### Genetic dissection of the BCL11A and HBS1L-MYB loci

Both loci overlap erythroid enhancer elements for adjacent genes *BCL11A* and *MYB* (Figure 2). They contain multiple independent association signals, indicating the likely presence of several functionally active DNA variants interfering with enhancer function. Our fine-mapping strategy was directed at defining the haplotypes carrying HbF-boosting alleles and at identifying putative functional variants residing therein.

**Figure 2.**
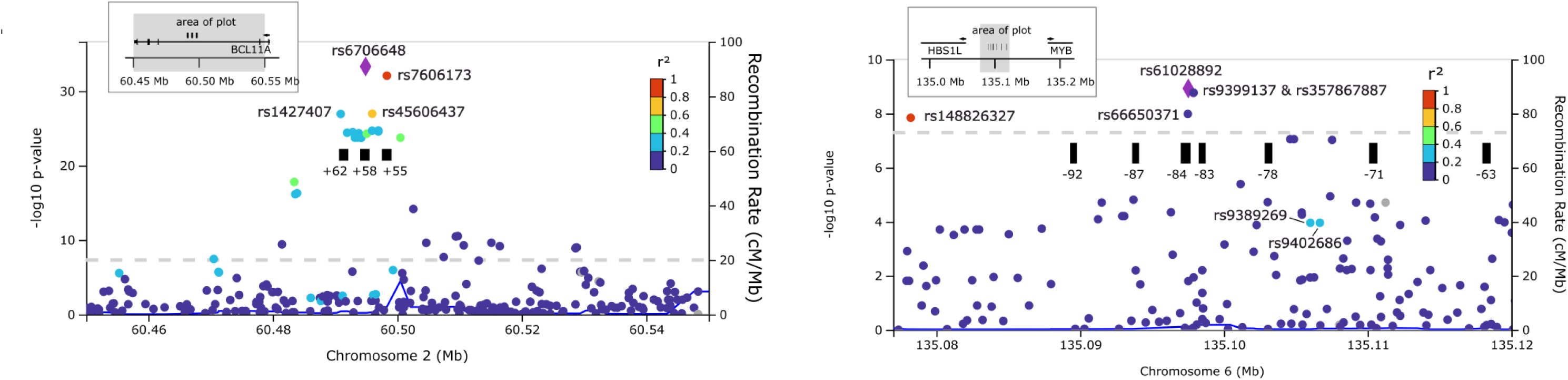
Regional association plots for *BCL11A* (left panel) and *HBS1L-MYB* loci (right panel). The dashed line shows the threshold for genome-wide significance (p < 5 x 10^-8^). The black rectangular blocks show key erythroid regulatory elements, detected as DNase I hypersensitive sites for *BCL11A* [29] and LDB1 binding sites for HBS1L-MYB [33].

First, we chose variants tagging HbF-relevant haplotypes aided by conditional association analysis. We then surveyed our phase-aligned genotype data to identify regional haplotypes defined by these variants, recorded their presence in patients, and measured their effects. To spotlight putative functional variants, we included functional sequence annotation in our fine-mapping using functional GCTA (fGCTA). To differentiate between variants in close linkage disequilibrium in African datasets that appeared to have a similar impact on HbF values, we performed additional association analysis in a European non-anaemic (TwinsUK) cohort.

#### BCL11A

Stepwise regression analysis initially yielded two partially independent signals, rs6706648 (our lead SNP at *BCL11A*) and rs1427407 (Table 2), a variant widely reported in other cohorts [15] that has been reported to disrupt critical transcription factor sites and is considered functional [32]. The fGCTA approach identified the same variants as in the standard conditional analysis (Table 2) for the erythroblast GATA1, K562, and HUDEP-2 annotations; rs1427407 overlapped with a GATA1 site (Supplementary Table 4, Supplementary Figure 2). When we conditioned on rs1427407 alone (Supplementary Table 5), the most significant SNP obtained was not rs6706648, but rs7565301, which is less frequent but with a similar allelic effect. To define haplotypes of interest, we also considered rs7606173, a widely reported SNP that we found in close LD with our lead SNP (D′ = 1, r^2^ = 0.91) (Figure 2). Accordingly, the power to distinguish between these two SNPs in our patient dataset is limited. Somewhat surprisingly, the LD between them is significantly less tight in European populations (EUR: D′ = 0.99, r^2^ = 0.61 versus YRI: D′ = 1.0, r^2^ = 0.91 in 1000 Genomes Phase III data implemented in LDlink [33]. Therefore, we decided to test both for HbF association (ln[%F-cells] trait) in the TwinsUK dataset and found rs7606173 to be substantially stronger (β = −0.296, p = 2.29 x 10^-79^ vs. β = -− 0.243, p = 1.27 x 10^-47^ of rs6706648) (Supplementary Table 6).

**Table 2.** Conditional analysis of chromosome 2 in the Nigerian cohort.

| Chr | SNP | Position | EA / EAF | Non-conditional |  | Conditional on rs6706648 |  | Conditional on rs6706648 + rs1427407 |  |
| --- | --- | --- | --- | --- | --- | --- | --- | --- | --- |
| | | | | β-value | p-value | β-value | p-value | β-value | p-value |
| 2 | rs6706648 | 60494905 | T / 0.41 | -0.395 | $4.96 \times 10^{-34}$ | - | - | - | - |
| 2 | rs1427407 | 60490908 | T / 0.25 | 0.41 | $1.14 \times 10^{-27}$ | 0.20 | $3.68 \times 10^{-7}$ | - | - |
| 2 | rs58789059 | 60509717 | A/0.17 | -0.28 | $3.03 \times 10^{-11}$ | -0.12 | 0.005 | -0.12 | 0.004 |

Chr: Chromosome; SNP: reference ID for the Single Nucleotide Polymorphism; EA: Effect allele; OA: Other allele; EAF: Effect allele frequency; $\beta$ : allelic effect size. Chromosomal positions are in hg38.

We used SNPs rs1427407, rs7565301, and rs7606173 to define haplotypes at *BCL11A*, of which we observed six, four of them with an allele frequency above 1% (Figure 3). We estimated the effect of the common *BCL11A* haplotypes on our trait (ln [HbF%]) compared to ‘basal’ haplotype 4, which contains low-HbF alleles for all three tag SNPs (‘GGC’). Haplotype 1 (‘**T**GG’), containing the high-HbF allele for rs1427407, had a significant trait-increasing effect (+0.537, p < 0.0001) as had haplotype 3 (‘G**A**G’), which contains the high-HbF allele for rs7565301 (+0.319, p < 0.0001). Haplotype 2 (‘GG**G**’), which contains the high-HbF allele only for rs7606173, had a smaller effect (+0.197), which was not significant, possibly due to its low frequency of 2.6%. Haplotypes 2 and 3 did not have significantly different effects. These HbF-boosting haplotype effects were validated in the UK SCD datasets (Supplementary Table 7).

**Figure 3.**
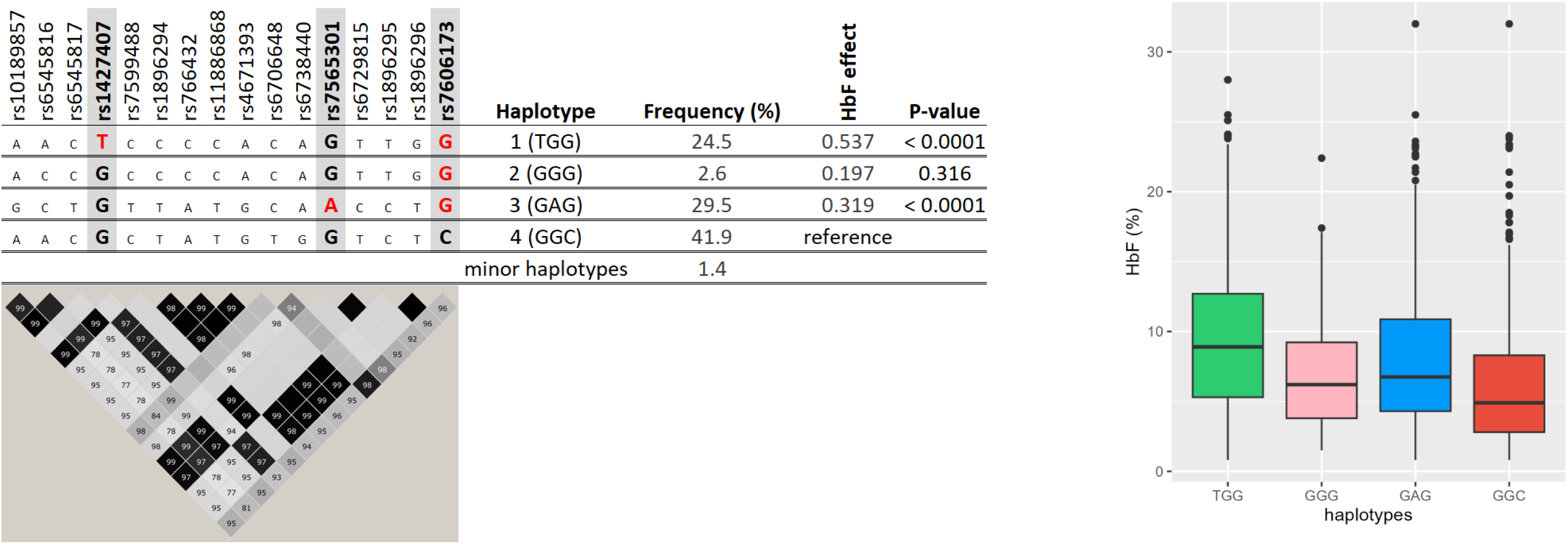
Major haplotypes at the *BCL11A* locus as defined by key markers rs1427407, rs7565301, and rs7606173. The left side of the graph shows the relationship of these tagging SNPs with other variants populating these haplotypes, i.e., associated alleles (top) and LD relationships (bottom). The right side shows haplotype effects on HbF values. LD plot: Each diamond shows the value of Dʹ and the LD indicates white (r^2^ = 0), shades of grey (0 < r^2^ < 1), and black (r^2^ = 1).

#### HBS1L-MYB

Conditional analysis detected two separate association signals, the lead SNP, rs61028892, and rs9399137 (Table 3). fGCTA analysis identified rs61028892 as the peak SNP following the conditional analysis (Table 2). The K562 annotation identified the same secondary signal (rs9399137) with the conditional analysis. However, erythroblast GATA1 and HUDEP-2 tracks identified a different secondary signal (rs66650371) (Supplementary Table 8, Supplementary Figure 3). All these SNPs overlapped with their respective annotations. The ‘basal’ haplotype 3 (‘GT’), (Figure 4), carrying low-HbF variants at both positions, was overwhelmingly frequent (94%), as it is typical for African populations [34]. We observed two other haplotypes: haplotype 1 (‘G**C**’, carrying the high-HbF allele at rs9399137) with an HbF increasing effect of +0.497 (p < 0.0001) and haplotype 2 (‘**C**T’, carrying the high-HbF allele at rs61028892) with an effect of +0.671 (p < 0.0001). Similar haplotype effects were confirmed in the UK SCD datasets as shown in the Supplementary Table 9.

**Figure 4.**
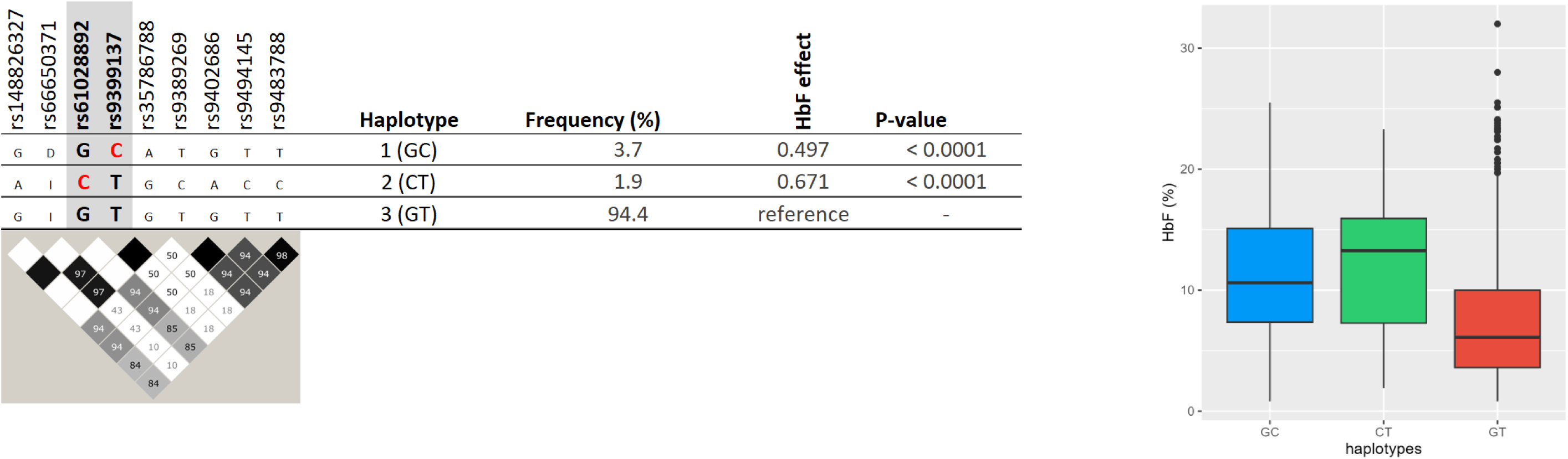
Major haplotypes at the HBS1L-MYB locus as defined by key markers rs61028892 and rs9399137. The left side of the graph shows the relationship of these tagging SNPs with other variants populating these haplotypes, i.e., associated alleles (top) and LD relationships (bottom). The right side shows haplotype effects on HbF values. LD plot: Each diamond shows the value of Dʹ and the LD indicates white (r^2^ = 0), shades of grey (0 < r^2^ < 1), and black (r^2^ = 1).

**Table 3.** Conditional analysis of chromosome 6 in the Nigerian cohort.

| Chr | SNP | Position | EA / EAF | Non-conditional |  | Conditional on rs61028892 |  | Conditional on rs61028892 + rs9399137 |  |
| --- | --- | --- | --- | --- | --- | --- | --- | --- | --- |
| | | | | β-value | p-value | β-value | p-value | β-value | p-value |
| 6 | rs61028892 | 135097526 | C/0.02 | 0.73 | $1.18 \times 10^{-9}$ | - | - | - | - |
| 6 | rs9399137 | 135097880 | C/0.04 | 0.50 | $1.71 \times 10^{-9}$ | 0.52 | $9.48 \times 10^{-10}$ | - | - |
| 6 | rs1135205 | 134960657 | C/0.25 | 0.12 | 0.00079 | 0.12 | 0.00087 | 0.14 | 0.00013 |

Chr: Chromosome; SNP: reference ID for the Single Nucleotide Polymorphism; EA: Effect allele; OA: Other allele; EAF: Effect allele frequency; $\beta$ : allelic effect size. Chromosomal positions are in hg38.

### Trait variability explained

We applied a multiple linear regression modelling to estimate the genetic effects on HbF (ln [HbF%]) using eight representative SNPs on *BCL11A* (rs7606173, rs1427407, rs6706648, rs6738440, rs7565301*) and HBS1L-MYB (*rs9399137, rs66650371 and rs61028892) and found that rs7606173, rs1427407, rs6738440, rs9399137, and rs61028892 account for 24.1% HbF variance in SCD patients.

### Search for functionally active genetic variants

We detected haplotypes with significant trait-increasing effects (Figure 3) at *BCL11A* (haplotypes 1 and 3) and two at *HBS1L-MYB* (haplotypes 1 and 2, Figure 4), each expected to harbour at least one functional DNA variant causing higher HbF levels in patients. At *BCL11A*, the tag SNP for haplotype 1 (rs1427407) itself is likely to be functional [32]. For haplotype 3, candidates are the tag SNP rs7565301, variants in close LD with it (rs6729815: D′ = 1.0, r^2^ = 1.0; rs7599488: D′ = 1.0, r^2^ = 1.0; rs6545817: D′ = 0.98, r^2^ = 0.96; rs10189857: D′ = 1.0, r^2^ = 0.98; rs6545816: D′ = 0.98, r^2^ = 0.73) or other, so far undetected variants. At *HBS1L-MYB*, a 3-bp deletion (rs66650371) in close LD (D′ = 1.0, r^2^ = 1.0) with the tag SNP for haplotype 1 (rs9399137) has been suggested to be functional [35, 36]. For haplotype 2, candidates are the tag SNP rs61028892, variants in close LD with it (rs148826327: D′ = 1.0, r^2^ = 1.0, rs116460276: r^2^ = D′ = 1.0, r^2^ = 1.0) or an undetected variant.

### Evaluation of novel association signals

A list of all genome-wide and suggestive loci detected in the Nigerian cohort is presented in Supplementary Table 10. The regional association plots for the novel genome-wide significant loci are shown in the Supplementary Figures 4-6.

For any novel locus detected near (above or below) the acknowledged threshold for genome-wide significance there is a chance of it being either a true (caused by functional DNA sequence changes) or a spurious association signal. To distinguish these scenarios, we sought to confirm (‘replicate’) our three novel signals in three further sickle cell populations of African descent, combined through meta-analysis (Table 1). We detected no significant association but the longitudinal data of 326 SCD African-American patients from St Jude Children’s Research Hospital showed a replication for *PIEZO2* (p = 0.00028) (Supplementary Table 11).

To search for additional association signals, we conducted further meta-analysis, involving the genome-wide association results of all 3582 patients (Nigeria, UK, Tanzania, and African-American). We identified one genome-wide significant locus and nine suggestive genome-wide significant loci, in addition to those detected in the Nigerian cohort alone (Figure 5, Supplementary Table 12).

**Figure 5.**
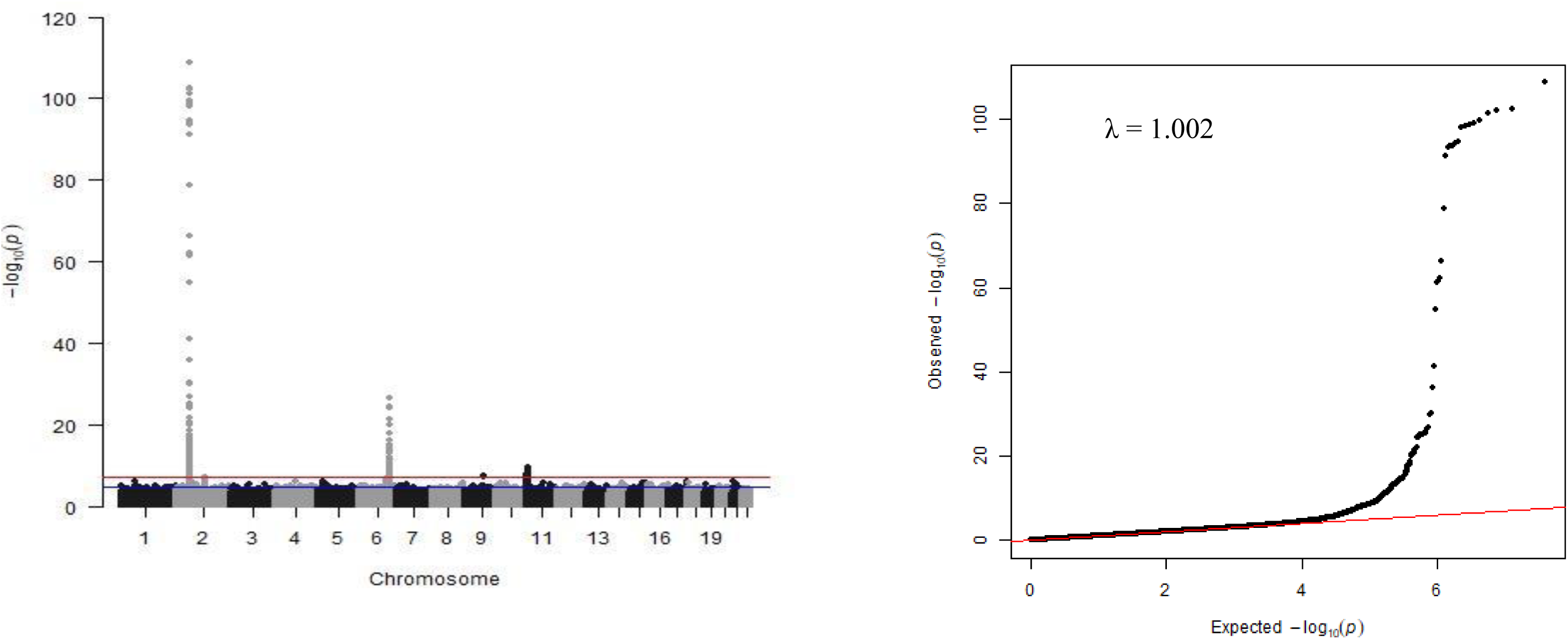
(a) Manhattan plot showing meta-analysis of 3582 SCD patients. The y-axis indicates the −log(*P*-value) for each variant in the meta-analysis and x-axis shows chromosomal position. (b). Quantile-Quantile plot with the genomic control inflation (λ) of 1.002.

## Discussion

We conducted a genome-wide quantitative-trait association study for fetal-haemoglobin levels in 1,006 Nigerian patients with sickle cell disease recruited from three regions of the country. At the genome-wide significance level, we detected two of the three known major loci for HbF persistence [7, 9, 37], *BCL11A* (chromosome 2) and *HBS1L-MYB* (*HMIP*) (chromosome 6), and three new loci, *SLC28A3* on chromosome 9, *TICRR* on chromosome 15, and *PIEZO2* on chromosome 18.

HbF persistence is a widely studied genetic trait in sickle cell patients due to its important disease-ameliorating and life-prolonging effect, the broad availability of a reliable assay system (HPLC), and its reduced complexity compared to other SCD-severity related traits. HbF reactivation is a key therapeutic goal, targeted not only by the disease-modifying drug, hydroxyurea, but is also the subject of promising gene therapy approaches presently undergoing clinical trials [38, 39]. *BCL11A* and *HBS1L-MYB* are major HbF loci present in all human populations, including in SCD patients [7, 9, 37], but the third, *Xmn1-HBG2*, residing within the beta-globin gene cluster, is detected only in patients where so-called ‘Senegal’ and ‘Arab-Indian’ haplotypes surrounding the sickle mutation are prevalent. While it was unsurprising to encounter *BCL11A* and *HBS1L-MYB* in the current study, the genetic dissection of these loci has provided us with candidates for the underlying functionally active sequence variants for biological experiments to study locus effects and mechanisms of action.

*BCL11A* (B-cell lymphoma/leukaemia 11A) is a known repressor of γ-globin expression. It participates in the haemoglobin switch from γ to β-globin, but genetic variants present in intron 2 of the gene are associated with moderate HbF persistence, detected as increased F-cell numbers and overall increased HbF levels. These variants occupy the erythroid enhancer region, characterised by three DNase I-hypersensitive sites (DHSs) situated at +55 kb, +58 kb, and +62 kb from the transcription start site (TSS)[32].

We detected two haplotypes (defined by rs1427407, rs7565301, and rs7606173) with strong HbF-increasing effect at this locus. The first (haplotype TGG) carries HbF-increasing alleles at rs1427407 (‘T’) and rs7606173 (‘G’). rs1427407 is located within GATA1 and TAL1 binding motifs at the +62 kb site, suggesting that the T allele may disrupt GATA1 binding. Thus, rs1427407-‘T’ is the likely causal variant driving the effect of this haplotype. A second haplotype with a significant HbF-increasing effect (haplotype 3) is defined by the presence of the rs7565301 variant (‘A’ allele) and rs7606173 (‘G’). Haplotype 2, carrying only rs7606173 (‘G’) but not rs7565301-‘A’, did not have a significant effect. Hence, we suggest that a causative sequence variant closely linked with rs7565301 is likely driving the significant effect of haplotype 3 and possibly also a weak (diluted) effect at haplotype 2. We presently cannot exclude that rs7606173 (‘G’) or a closely linked variant cause the HbF- raising effects of haplotypes 1, 2, and 3.

Association of the *BCL11A* locus with HbF has been previously reported in diverse populations with different SNPs on *BCL11A* showing the strongest association with HbF levels in SCD patients (Supplementary Table 3) [7, 8, 15, 40]. In our dataset, these reported SNPs map to the same effect haplotypes we have identified. Thus, we suggest that the same underlying causative variants are responsible in all non-anaemic and patient populations studied so far.

The Tanzanian SCD cohort and the African-American cohort of Cooperative Study of SCD (CSSCD) showed rs1427407 as the strongest association signal with HbF, with rs6545816 and rs7606173 detected as secondary signals [15, 32]. The difference with our findings underscores the influence of population-specific allele frequencies and LD patterns on the identity of lead (sentinel) SNPs reported in specific studies.

At *HBS1L-MYB*, we detected two haplotypes (defined by rs61028892 and rs9399137) with significant HbF-boosting effect. Haplotype 1, tagged by rs9399137-‘C’, causes what has previously been described as the *HMIP-2A* sub-locus [34]. It harbours a 3-bp deletion (rs66650371), which was proposed to be a functional DNA change [35] and was found to disrupt a TAL-1 binding motif within a critical enhancer element for *MYB*, the gene encoding an essential transcriptional regulator of erythropoiesis [36]. Decreased promoter interaction (looping) of an allele carrying the deletion was observed in K562 cells [36], but it is not clear whether this was due to the deletion itself or closely linked variants occupying the same haplotype. HbF-increasing haplotype 2 is defined by the presence of rs61028892-‘C’, which also carries two closely linked alleles rs148826327-‘A’ (D′ = 1.0, r^2^ = 1.0), and rs116460276- ‘G’ (D′ = 1.0, r^2^ = 1.0). It is also linked to a series of variants that have previously served to define the *HMIP-2B* sublocus [34, 41]. rs61028892 is situated at the same *MYB* enhancer element (‘−84 LDB1 site’) as the 3-bp deletion rs66650371 and has previously been proposed as an additional casual variant in the *HMIP* region [42]. This suggests that the HMIP-2 locus might not contain sub-loci, but three functional alleles affecting enhancer element -84, two increasing HbF (rs66650371-del and rs61028892-‘C’) and one representing the baseline, low-HbF state (haplotype ‘**GT**’). The more precise definition of the *HMIP-2* locus we have obtained with these data highlights the significantly lower high-HbF allele frequency at this site in African populations (5.6% total haplotype frequency) compared to individuals of European descent (>20%) and or to patients with substantial European admixture [7, 43].

Consistent with previous studies, HbF variability explained by *BCL11A* and *HMIP* in our study is 24.1%, which is lower than in non-anaemic populations (∼ 45%) [7, 12, 13]. Sex and age (up to 3%) are only minor contributors to the overall trait variability. Thus, after major HbF loci are taken into consideration, a substantial number of loci with smaller effects or loci that are sickle-cell specific remain to be discovered.

Lastly, the three novel HbF loci detected, *SLC28A3* on chromosome 9 and *TICRR* on chromosome 15 with genome-wide significance, and *PIEZO2* on chromosome 18 close to the acknowledged threshold were not confirmed in the British, Tanzanian, and African-American patients. Thus, whether these new loci harbour functional, HbF-affecting variants or represent spurious association signals will have to be resolved through additional genetic or biological investigation.

## Conclusions

In the current study, we have refined the definition of two major fetal-haemoglobin persistence loci, *BCL11A* and *HS1L-MYB*, which will aid future functional studies that seek to identify new ways to induce HbF expression in patients with sickle cell disease and beta thalassaemia. We have detected new putative HbF loci, which are presently unconfirmed.

Finding biological pathways influencing SCD severity remains a major goal for genetic studies with patient populations. The statistical power to detect loci with small effects lies in the assembly of large patient cohorts and in combining world-wide studies.

## Data Availability

All data produced in the present study are available upon reasonable request to the authors.

## List of Abbreviations

*BCL11A*: B-cell lymphoma/leukemia 11A
CSSCD: African-American cohort of Cooperative Study of SCD
DHS: DNase I-hypersensitive site
fGCTA: functional GCTA
*GCTA*: *G*enome-wide complex trait analysis
HbF: Fetal haemoglobin
HbS: haemoglobin S
*HBS1L-MYB*: HBS1 like Translational GTPase-MYB Proto-Oncogene
HbSS: Homozygosity for HbS
HbSβ: Compound heterozygote state for HbS and beta thalassaemia (HbSβ^0^/ HbSβ^+^)
*HMIP*: HBS1L-MYB intergenic polymorphism
LD: Linkage disequilibrium
MAF: Minor Allele Frequency
SCD: sickle cell disease
TOPMed: Trans-Omics for Precision Medicine
TSS: transcription start site.

## Acknowledgements

The authors thank the Sickle Cell Foundation Nigeria, Lagos, Nigeria, and Dr Annette Akinsete for providing an enabling environment and facility for patients’ recruitment. We are grateful to all the patients for their participation in this study. We acknowledge Mr. Musa Mohammed who coordinated the laboratory activities including blood sampling and haematological investigations at Ahmadu Bello University Teaching Hospital, Zaria, Nigeria. The authors are also grateful to Prof Lawal W. Umar, the Coordinator of Institute of Child Health, (ICH) Banzazzau, Zaria, Nigeria; and the Resident doctors, house officers, and all staff at the Paediatric Haematology/Oncology Unit, Ahmadu Bello University Teaching Hospital, Zaria, Nigeria.

## Funding

This work was funded in principle by MRC grant MR/T013389/1 (to SM and JS), with additional funding from EPSRC (Newton/GCRF) grant EP/X527920/1, from LIBRA and from the King’s College Hospital Charity. Twins UK is supported by the Wellcome Trust Core Award Grant Number 203141/Z/16/Z with additional support from the NIHR Oxford BRC. AG and CG were supported by the Wellcome Trust Core Award Grant Number 203141/Z/16/Z with additional support from the NIHR Oxford BRC. The views expressed are those of the author(s) and not necessarily those of the NHS, the NIHR or the Department of Health.

## Author’s contributions

Conception and design: SM, JS, OO, TA, ON. Patient recruitment: TA, AIO, BI, KG, JB, HI, UA, RC, JAF, BYJ, NMA, AH, BI, JM, HA, ON. Sample preparation: OO, AA, BI, KG, HR, RE, SN; Data curation: OO, RC, HP, S-HL, SR, GK, KG, RS, LM; Data Analysis and interpretation: OO, KG, BSM, SR, GK, CG, AG; Supervision: SM, TA, HA, SN, ON, IAU; Manuscript writing: OO and SM with input from all authors. All authors read and approved the final manuscript. Coordinated the project: SM and JS.

## Availability of data and materials

The data that support the findings of this study are available from the corresponding author upon reasonable request.

## Ethics approval and consent to participate

Written informed consent was obtained from the patients and when children were recruited, their parents/guardians signed the consent form while children above the age of seven gave assent. This study was approved by the Health Research Ethics Committee of Lagos University Teaching Hospital (ADM/DCST/HREC/1686), University of Abuja Teaching Hospital (FCT/UATH/HREC/PR/144), and Ahmadu Bello University ethics committee (ABUCUHSR/2021/031). This study was conducted in accordance with the Helsinki Declaration (1975, as revised in 2008).

## Consent for publication

Not Applicable

## Competing interests

The authors declare that they have no competing interests.

## Notes

### Competing Interest Statement

The authors have declared no competing interest.

### Author Declarations

This study was approved by the Health Research Ethics Committee of Lagos University Teaching Hospital (ADM/DCST/HREC/1686), University of Abuja Teaching Hospital (FCT/UATH/HREC/PR/144), and Ahmadu Bello University ethics committee (ABUCUHSR/2021/031).

