## Supplemental Tables and Figures for "The genetic dissection of fetal haemoglobin persistence in sickle cell disease in Nigeria"

**Supplementary Table 1. Characteristics of the Nigerian cohort included in the GWAS**

| | | | Age (years) | | | HbF (%) | | |
|---|---|---|---|---|---|---|---|---|
| | **N** | **Sex (M/F)** | **Median [IQR]** | **Min** | **Max** | **Median [IQR]** | **Min** | **Max** |
| ABUJA | 145 | 64/81 | 16[12-22] | 5 | 41 | 9.4 [5.80 – 13.90] | 0.8 | 28.0 |
| LAGOS | 638 | 314/324 | 14 [9 -23] | 5 | 60 | 5.8 [3.40 – 9.30][a] | 0.8 | 32.0 |
| ZARIA | 223 | 97/126 | 18 [10-23] | 5 | 45 | 6.1 [4.00 – 10.50][b] | 0.8 | 23.3 |
| **TOTAL** | **1006** | **475/531** | **15 [9 -23]** | **5** | **60** | **6.69 [3.67 – 11.02]** | **0.8** | **32.0** |

a: Significantly lower HbF levels compared to patients in ABUJA site ($P$ = 1.77 x $10^{-9}$).
b: Significantly lower HbF levels compared to patients in ABUJA site ($P$ = 5.1 x $10^{-5}$).

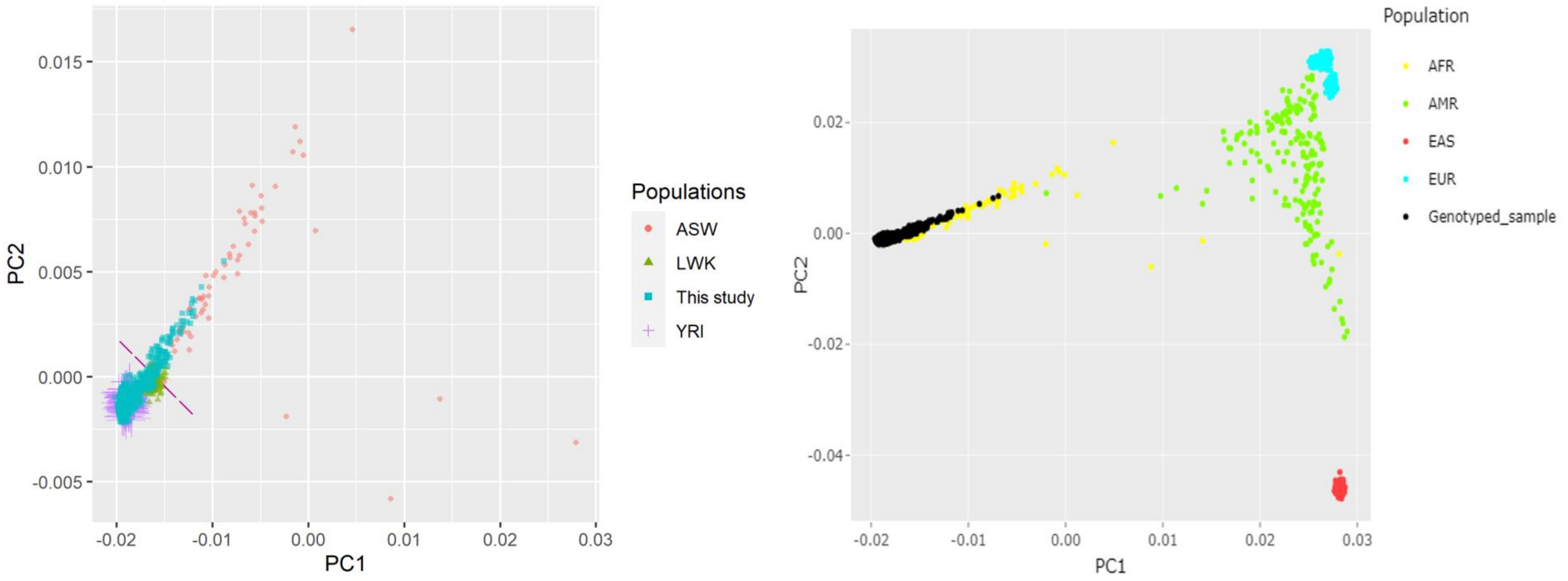


Supplementary Figure 1. (a) The pricincipal component analysis shows individuals from the Southern Nigeria (below the dashed line) and those from the Northen Nigeria (above the dashed line). YRI: Yoruba in Ibadan population; LWK: Luhya in Webuye, Kenya; ASW: Americans of African Ancestry in Southwest USA. (b). The principal component analysis of the Nigerian cohort alongside the major population ancestries. AFR: Africa; AMR: America; EAS: East Asia; EUR: Europe.

Supplementary Table 2. Quality control summary

| QC step | Individuals excluded | SNPs excluded |
|---|---|---|
| **Per-individual QC** | | |
| Genotype call rate < 90%* | 77 | |
| Sex discrepancy samples | 21 | |
| Heterozygosity outliers | 20 | |
| Genetically identical samples | 20 | |
| **SBeta+ | 6 | |
| **Patients < 5 years | 8 | |
| Remaining individuals: **1006** | | |
| **Per-marker QC** | | |
| SNP call rate < 97% | | 61,878 |
| MAF < 1% | | 266,727 |
| HWE $p < 10^{-8}$ | | 5616 SNPs |
| Non-autosomal SNPs | | 44,294 |
| Remaining SNPs: **1,925,391** | | |

*Samples with genotype call rates < 90% were excluded from the Illumina Genome Studio software following our previously published GWAS quality control protocol. Samples excluded due to heterozygosity outliers were mainly from the region where consanguinity is common. #: Of these, 50941 SNPs were zeroed during pre-QC steps in the Genome studio.

**Supplementary Table 3. Previous HbF-associated genome-wide significant SNPs in three major loci**

| Chr/Gene | Top SNP | MAF | p-value | LD (D', $r^2$) | Sample size | Population | Cohort | Study type | Authors |
|---|---|---|---|---|---|---|---|---|---|
| Chr 2: BCL11A | | | | | | | | | |
| | rs1427407 | 0.14 | $2.5 \times 10^{-20}$ | 0.94, 0.16 | 179 | Northern Europeans | Twins UK | GWAS | [1] |
| | rs4671393 | 0.27 | $2.0 \times 10^{-42}$ | 1.0, 0.23 | 1275 | African-American | CSSCD | GWAS | [2] |
| | rs766432 | 0.28 | $2.61 \times 10^{-21}$ | 1.0, 0.23 | 848 | African-American | CSSCD | GWAS | [3] |
| | rs766432 | 0.276 | $5.36 \times 10^{-58}$ | 1.0, 0.23 | 2040 | African-American | Multicentre-study | Meta-analysis | [4] |
| | rs7606173 | 0.45 | $5.14 \times 10^{-16}$ | 1.0, 0.91 | 440 | African_American | SIT Trial | GWAS | [5] |
| | rs1427407 | 0.22 | $3.74 \times 10^{-53}$ | 0.94, 0.16 | 1213 | East-Africa | Tanzania | GWAS | [6] |
| | rs1896295 | 0.25 | $2.49 \times 10^{-26}$ | 1.0, 0.22 | 585 | African-American | BCM & SCCRIP | GWAS | [7] |
| | rs6706648 | 0.41 | $4.96 \times 10^{-34}$ | | 1006 | West-Africa | Nigeria | GWAS | Current study |

| | | | | | | | | | |
|---|---|---|---|---|---|---|---|---|---|
| Chr 6: HBS1L-MYB | | | | | | | | | |
| | rs9399137 | 0.23 | $2.8 \times 10^{-27}$ | 1.0, 0.001 | 179 | Northern Europeans | Twins UK study | GWAS | [1] |
| | rs9399137 | 0.06 | $5.0 \times 10^{-11}$ | 1.0, 0.001 | 1275 | African-American | CSSCD | GWAS | [2] |
| | rs9494145 | 0.07 | $4.32 \times 10^{-17}$ | 1.0, 0.59 | 2040 | African-American | Multicentre | Meta-analysis | [4] |
| | rs9494145 | 0.05 | $2.42 \times 10^{-10}$ | 1.0, 0.59 | 1213 | East-Africa | Tanzania | GWAS | [6] |
| | rs116460276 | 0.02 | $3.06 \times 10^{-10}$ | 1.0, 1.0 | 585 | African-American | SIT trial | GWAS | [7] |
| | rs61028892 | 0.02 | $1.18 \times 10^{-9}$ | | 1006 | West-African | Nigeria | GWAS | Current study |
| Chr 11 | | | | | | | | | |
| *XmnI-HBG2* | rs7482144 | 0.33 | $2.0 \times 10^{-30}$ | - | 179 | Northern Europeans | Twins UK study | GWAS | [1] |
| *XmnI-HBG2* | rs7482144 | 0.07 | $4.0 \times 10^{-7}$ | - | 1275 | African-American | CSSCD | GWAS | [2] |
| *OR51B5/OR51B6* | rs5006884 | 0.13 | $4.73 \times 10^{-8}$ | - | 848 | African-American | CSSCD | GWAS | [3] |

LD (D’, $r^2$) relationship between our lead SNP and SNPs reported in previous studies using LDlink: YRI (Yoruba in Ibadan, Nigeria) population in 1000 Genome Project[8]. CSSCD: Cooperative Study of SCD; BCM: Baylor College of Medicine cohort; SCCRIP: St. Jude Children’ Research Hospital Sickle Cell Clinical Research and Intervention Program; SIT: Silent Infarct Transfusion.

**Supplementary Table 4: fGCTA results for the *BCL11A* locus**

| | | | | | Scaled Summary Statistics | | | COJO output | | | |
|---|---|---|---|---|---|---|---|---|---|---|---|
| **Annotation** | **SNP** | **Chr** | **Position** | **Frequency** | **b** | **se** | **p-value** | **bJ** | **bJ_se** | **pJ** | **Overlap with annotation** |
| GATA1 | rs6706648* | 2 | 60722040 | 0.413 | -0.395 | 0.033 | 1.24 E-33 | -0.288 | 0.039 | 1.32E-13 | FALSE |
| GATA1 | rs1427407 | 2 | 60718043 | 0.253 | 0.409 | 0.037 | 4.57E-28 | 0.257 | 0.044 | 5.88E-09 | TRUE |
| HUDEP-2 | rs6706648* | 2 | 60722040 | 0.413 | -0.395 | 0.033 | 1.24E-33 | -0.292 | 0.039 | 4.98E-14 | FALSE |
| HUDEP-2 | rs1427407 | 2 | 60718043 | 0.253 | 0.409 | 0.038 | 2.86E-27 | 0.254 | 0.045 | 1.21E-08 | FALSE |
| K562 | rs6706648* | 2 | 60722040 | 0.413 | -0.395 | 0.033 | 1.24E-33 | -0.292 | 0.039 | 4.89E-14 | FALSE |
| K562 | rs1427407 | 2 | 60718043 | 0.253 | 0.409 | 0.038 | 2.86E-27 | 0.254 | 0.045 | 1.20E-08 | FALSE |

GCTA-COJO p-value threshold: $1x10^{-5}$, fGCTA enrichment parameter: x2.5; * Peak SNP for this locus. Position: Chromosomal positions are in hg37; GATA1: GATA1 ChIP-seq on human peripheral derived blood-erythroblast; b = effect size, SE= standard error, p-value from the scaled summary statistics; bJ = effect size, sbJ_se= standard error and pJ = p-value from a joint analysis of all the selected SNPs.

**Supplementary Table 5. List of SNPs obtained after conditional analysis on *rs1427407*.**

| Chr | rsID | bp | EA | freq | β | se | p | βC | βC_se | pC |
|---|---|---|---|---|---|---|---|---|---|---|
| 2 | rs7565301 | 60496131 | A | 0.30169 | 0.104313 | 0.034342 | 0.002386 | 0.22691 | 0.034482 | 4.69E-11 |
| 2 | rs6706648 | 60494905 | T | 0.412525 | -0.3947 | 0.032454 | 4.96E-34 | -0.23121 | 0.035183 | 4.97E-11 |
| 2 | rs7606173 | 60498316 | C | 0.434292 | -0.39256 | 0.032912 | 8.50E-33 | -0.2344 | 0.035687 | 5.09E-11 |
| 2 | rs6729815 | 60496537 | C | 0.307157 | 0.105949 | 0.034167 | 0.001929 | 0.224669 | 0.034313 | 5.84E-11 |
| 2 | rs7599488 | 60491212 | T | 0.303181 | 0.101711 | 0.034315 | 0.003036 | 0.224745 | 0.034448 | 6.83E-11 |
| 2 | rs6545817 | 60488044 | T | 0.307271 | 0.095021 | 0.034126 | 0.005362 | 0.219086 | 0.03424 | 1.57E-10 |
| 2 | rs6738440 | 60495106 | G | 0.27998 | -0.36236 | 0.035081 | 5.20E-25 | -0.23657 | 0.037066 | 1.75E-10 |
| 2 | rs6709302 | 60500494 | A | 0.313347 | -0.356 | 0.034868 | 1.79E-24 | -0.23542 | 0.036905 | 1.78E-10 |
| 2 | rs10189857 | 60486100 | G | 0.307654 | 0.093908 | 0.034117 | 0.005913 | 0.217889 | 0.034228 | 1.94E-10 |
| 2 | rs13019832 | 60483436 | A | 0.428926 | -0.28073 | 0.031961 | 1.58E-18 | -0.20951 | 0.033245 | 2.93E-10 |
| 2 | rs45606437 | 60495973 | A | 0.341182 | -0.36801 | 0.033736 | 1.05E-27 | -0.22505 | 0.035963 | 3.90E-10 |
| 2 | rs6545816 | 60487726 | C | 0.340457 | 0.079935 | 0.033026 | 0.015506 | 0.207082 | 0.033104 | 3.96E-10 |

Here we showed only p-values ≤ $10^{-10}$. EA: effect allele; β = effect size; se: standard error; p= pvalue; βC = effect size after conditioning on rs1427407; βC_se: standard error after conditioning on rs1427407; pC = p-value after conditioning on rs1427407.

**Supplementary Table 6. List of selected *BCL11A* SNPs from the 5060 UK Twins**

| Chr | SNP | bp | A1 | A2 | Freq | b | se | p |
|---|---|---|---|---|---|---|---|---|
| 2 | rs10189857 | 60486100 | G | A | 0.426581 | 0.043076 | 0.015578 | 0.00569 |
| 2 | rs6545816 | 60487726 | A | C | 0.456719 | 0.044903 | 0.015558 | 0.003899 |
| 2 | rs6545817 | 60488044 | C | T | 0.458004 | 0.045101 | 0.015567 | 0.003766 |
| 2 | rs1427407 | 60490908 | T | G | 0.152569 | 0.501033 | 0.021644 | 1.49E-118 |
| 2 | rs7599488 | 60491212 | T | C | 0.425 | 0.044047 | 0.01555 | 0.004616 |
| 2 | rs1896294 | 60491939 | C | T | 0.298913 | 0.331639 | 0.016968 | 4.59E-85 |
| 2 | rs766432 | 60492835 | C | A | 0.145455 | 0.505857 | 0.022067 | 2.72E-116 |
| 2 | rs11886868 | 60493111 | C | T | 0.299012 | 0.330788 | 0.016955 | 9.07E-85 |
| 2 | rs4671393 | 60493816 | A | G | 0.144763 | 0.505039 | 0.022111 | 1.80E-115 |
| 2 | rs6706648 | 60494905 | T | C | 0.319071 | -0.24288 | 0.016754 | 1.27E-47 |
| 2 | rs6738440 | 60495106 | G | A | 0.288636 | -0.20583 | 0.017407 | 2.90E-32 |
| 2 | rs7565301 | 60496131 | A | G | 0.266897 | 0.020675 | 0.017494 | 0.237272 |
| 2 | rs6729815 | 60496537 | T | C | 0.463933 | 0.041718 | 0.015478 | 0.007031 |
| 2 | rs1896295 | 60496951 | T | C | 0.144664 | 0.504048 | 0.022052 | 1.25E-115 |
| 2 | rs1896296 | 60496952 | G | T | 0.144565 | 0.503899 | 0.022043 | 1.17E-115 |
| 2 | rs7606173 | 60498316 | C | G | 0.435474 | -0.29618 | 0.015701 | 2.29E-79 |

A1: effect allele; A2: other allele; b = effect size; se: standard error; p = p-value; SNPs in grey refer to rs6706648 and rs7606173 that were investigated for fine-mapping in the white-British Twins' cohort.

**Supplementary Table 7.** Haplotype association analysis of *BCL11A* SNPs (rs1427407-rs7565301-rs7606173) in the UK SCD patients

| Haplotype | Frequency | HbF effect | P-value |
|---|---|---|---|
| TGG | 25.3 | 0.561 | < 0.001 |
| GGG | 3.5 | 0.122 | 0.844 |
| GAG | 29.3 | 0.213 | < 0.001 |
| GGC | 40.7 | reference | |

**Supplementary Table 8:** fGCTA results for the *HMIP* locus

| | | | | | **Scaled Summary Statistics** | | | **COJO output** | | | |
|---|---|---|---|---|---|---|---|---|---|---|---|
| **Annotation** | **SNP** | **Chr** | **Position_b37** | **Frequency** | **b** | **se** | **p-value** | bJ | bJ_se | pJ | **Overlap with annotation** |
| **GATA1** | rs61028892* | **6** | 135418664 | **0.018** | **0.732** | **0.118** | **4.73E-10** | 0.753 | 0.119 | 2.18E-10 | TRUE |
| **GATA1** | rs66650371 | **6** | 135418632 | **0.032** | **0.514** | **0.087** | **4.12E-09** | 0.531 | 0.088 | 1.74E-09 | TRUE |
| HUDEP-2 | rs61028892* | 6 | 135418664 | 0.018 | 0.732 | 0.118 | 4.73E-10 | 0.753 | 0.119 | 2.14E-10 | TRUE |
| HUDEP-2 | rs66650371 | 6 | 135418632 | 0.032 | 0.514 | 0.087 | 4.12E-09 | 0.531 | 0.088 | 1.72E-09 | TRUE |
| K562 | rs61028892* | 6 | 135418664 | 0.018 | 0.732 | 0.118 | 4.73E-10 | 0.756 | 0.119 | 2.14E-10 | TRUE |
| K562 | rs9399137 | 6 | 135419018 | 0.037 | 0.504 | 0.082 | 6.83E-10 | 0.521 | 0.083 | 3.01E-10 | TRUE |

GCTA-COJO p-value threshold: $1x10^{-5}$, fGCTA enrichment parameter: x2.5; * Peak SNP for this locus. Position: Chromosomal positions are in hg37; GATA1: GATA1 ChIP-seq on human peripheral derived blood-erythroblast; b = effect size, SE= standard error, p-value from the scaled summary statistics; bJ = effect size, sbJ_se= standard error and pJ = p-value from a joint analysis of all the selected SNPs.

**Supplementary Table 9.** Haplotype association analysis of *HBS1L-MYB* (rs61028892-rs9399137) in the UK SCD patients

| Haplotype | Frequency | HbF effect | P-value |
|---|---|---|---|
| GC | 5.0 | 0.486 | < 0.001 |
| CT | 1.6 | 0.892 | < 0.001 |
| GT | 93.5 | reference | |

**Supplementary Table 10. List of all suggestive and genome-wide significant SNPs**

| chr | name | bp | A1 | A2 | freq | b | se | p |
|---|---|---|---|---|---|---|---|---|
| 2 | rs6706648 | 60494905 | T | C | 0.412525 | -0.3947 | 0.032454 | 4.96E-34 |
| 2 | rs7606173 | 60498316 | C | G | 0.434292 | -0.39256 | 0.032912 | 8.50E-33 |
| 2 | rs45606437 | 60495973 | A | AC | 0.341182 | -0.36801 | 0.033736 | 1.05E-27 |
| 2 | rs1427407 | 60490908 | T | G | 0.252982 | 0.409466 | 0.037563 | 1.14E-27 |
| 2 | rs1896296 | 60496952 | G | T | 0.2715 | 0.38149 | 0.0366 | 1.95E-25 |
| 2 | rs11434093 | 60495961 | C | CA | 0.2695 | 0.383339 | 0.036793 | 2.04E-25 |
| 2 | rs1896295 | 60496951 | T | C | 0.271 | 0.381471 | 0.036691 | 2.56E-25 |
| 2 | rs766432 | 60492835 | C | A | 0.271869 | 0.37791 | 0.036424 | 3.21E-25 |
| 2 | rs1896294 | 60491939 | C | T | 0.271372 | 0.378659 | 0.036553 | 3.80E-25 |
| 2 | rs4671393 | 60493816 | A | G | 0.2735 | 0.377705 | 0.036513 | 4.44E-25 |
| 2 | rs6738440 | 60495106 | G | A | 0.27998 | -0.36236 | 0.035081 | 5.20E-25 |
| 2 | rs11886868 | 60493111 | C | T | 0.269384 | 0.379162 | 0.036752 | 5.92E-25 |
| 2 | rs10195871 | 60493454 | A | G | 0.273857 | 0.373505 | 0.036484 | 1.35E-24 |
| 2 | rs34211119 | 60493183 | GT | G | 0.275 | 0.373377 | 0.036562 | 1.75E-24 |
| 2 | rs10172646 | 60493622 | G | A | 0.275 | 0.373377 | 0.036562 | 1.75E-24 |
| 2 | rs6709302 | 60500494 | A | G | 0.313347 | -0.356 | 0.034868 | 1.79E-24 |
| 2 | rs7557939 | 60494212 | G | A | 0.274775 | 0.373549 | 0.036592 | 1.82E-24 |
| 2 | rs7584113 | 60494176 | A | G | 0.2745 | 0.37325 | 0.036571 | 1.86E-24 |
| 2 | rs13019832 | 60483436 | A | G | 0.428926 | -0.28073 | 0.031961 | 1.58E-18 |
| 2 | rs45484694 | 60483892 | CT | C | 0.230143 | 0.328752 | 0.039205 | 5.05E-17 |
| 2 | rs11692396 | 60483603 | G | A | 0.223161 | 0.327615 | 0.039257 | 7.10E-17 |
| 2 | rs72962585 | 60502567 | G | A | 0.220825 | -0.29849 | 0.038328 | 6.82E-15 |
| 2 | rs58789059 | 60509717 | A | G | 0.172465 | -0.27711 | 0.0417 | 3.03E-11 |
| 2 | rs7340264 | 60509394 | A | G | 0.185572 | -0.26581 | 0.040105 | 3.40E-11 |
| 2 | rs72962596 | 60515110 | T | C | 0.0996 | -0.33283 | 0.052535 | 2.37E-10 |
| 2 | rs72962586 | 60504631 | T | C | 0.101392 | -0.3265 | 0.051566 | 2.42E-10 |
| 2 | rs6732518 | 60481462 | C | T | 0.298387 | 0.226463 | 0.036187 | 3.90E-10 |
| 2 | rs72962592 | 60510666 | T | C | 0.163519 | -0.26563 | 0.042757 | 5.22E-10 |
| 2 | rs72962602 | 60516470 | G | A | 0.164343 | -0.26291 | 0.042616 | 6.86E-10 |
| 2 | rs555276704 | 60528808 | G | GTAA | 0.16633 | -0.26019 | 0.042609 | 1.02E-09 |
| 2 | rs72964419 | 60528663 | T | C | 0.160263 | -0.26403 | 0.043291 | 1.07E-09 |
| 2 | rs79059225 | 60528627 | T | C | 0.164813 | -0.25899 | 0.042683 | 1.30E-09 |
| 2 | rs66488669 | 60507448 | G | A | 0.230343 | -0.21291 | 0.037931 | 1.99E-08 |
| 2 | rs4672393 | 60470519 | A | C | 0.379602 | -0.18147 | 0.032941 | 3.61E-08 |
| 2 | rs114125602 | 60513084 | A | G | 0.027638 | -0.5223 | 0.096339 | 5.91E-08 |
| 2 | rs72964414 | 60523168 | T | C | 0.148851 | -0.21822 | 0.044088 | 7.44E-07 |
| 6 | rs61028892 | 135097526 | C | G | 0.017982 | 0.732037 | 0.120351 | 1.18E-09 |
| 6 | rs9399137 | 135097880 | C | T | 0.037276 | 0.503912 | 0.083658 | 1.71E-09 |
| 6 | rs35786788 | 135097904 | A | G | 0.037276 | 0.503912 | 0.083658 | 1.71E-09 |
| 6 | rs66650371 | 135097494 | T | TTAC | 0.032096 | 0.513867 | 0.089748 | 1.03E-08 |
| 6 | rs148826327 | 135078218 | A | G | 0.016 | 0.723182 | 0.127556 | 1.43E-08 |
| 6 | rs115099895 | 135104618 | A | G | 0.034328 | 0.464427 | 0.086831 | 8.86E-08 |
| 6 | rs114603312 | 135104880 | C | T | 0.034328 | 0.464427 | 0.086831 | 8.86E-08 |
| 6 | rs114398597 | 135107536 | G | A | 0.034363 | 0.463423 | 0.086834 | 9.45E-08 |

| | | | | | | | | |
|---|---|---|---|---|---|---|---|---|
| 9 | rs115555854 | 84339933 | A | C | 0.014955 | -0.72726 | 0.130523 | 2.52E-08 |
| 11 | rs1406381 | 62301213 | G | T | 0.320344 | -0.16572 | 0.03341 | 7.04E-07 |
| 14 | rs191465524 | 52721225 | G | T | 0.015936 | -0.63712 | 0.123409 | 2.44E-07 |
| 14 | rs188152509 | 52740738 | G | C | 0.01592 | -0.63708 | 0.123406 | 2.44E-07 |
| 15 | rs140496989 | 89619296 | A | G | 0.045817 | -0.42712 | 0.077342 | 3.34E-08 |
| 15 | rs78432130 | 89643261 | G | T | 0.045862 | -0.40726 | 0.077303 | 1.38E-07 |
| 15 | rs137993810 | 89628452 | G | C | 0.043327 | -0.41851 | 0.079446 | 1.38E-07 |
| 15 | rs78973372 | 89589437 | T | G | 0.045364 | -0.40542 | 0.077696 | 1.81E-07 |
| 15 | rs146039045 | 89616003 | A | G | 0.055224 | -0.3489 | 0.069713 | 5.59E-07 |
| 16 | rs188481235 | 1446101 | T | C | 0.019901 | -0.54714 | 0.111648 | 9.56E-07 |
| 18 | rs58817161 | 10818373 | C | T | 0.017396 | -0.63287 | 0.117935 | 8.04E-08 |
| 18 | rs142386135 | 10819008 | G | A | 0.017396 | -0.63287 | 0.117935 | 8.04E-08 |
| 18 | rs143311786 | 10820712 | G | A | 0.017396 | -0.63287 | 0.117935 | 8.04E-08 |

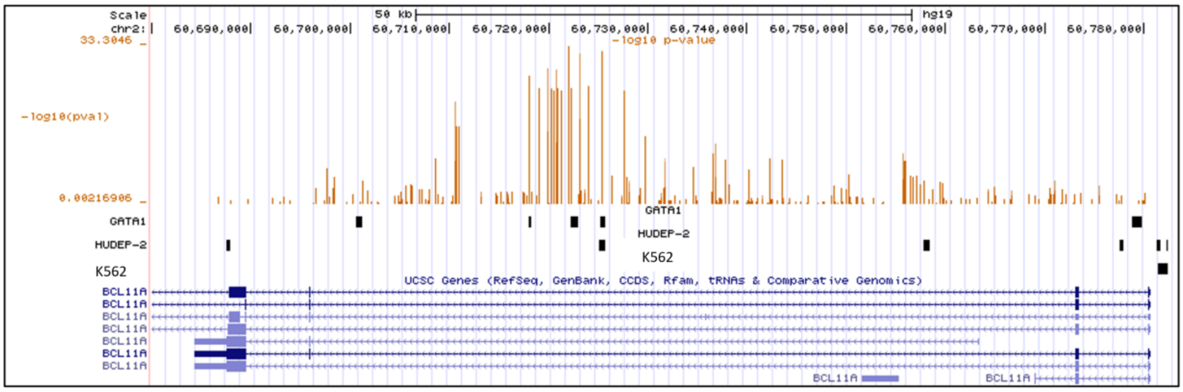


**Supplementary Figure 2:** UCSC plot for the *BCL11A* locus. Tracks in order: log10 p-values for variants; GATA1 ChIP-seq annotation; HUDEP-2 annotation; K562 annotation; genes in this locus.

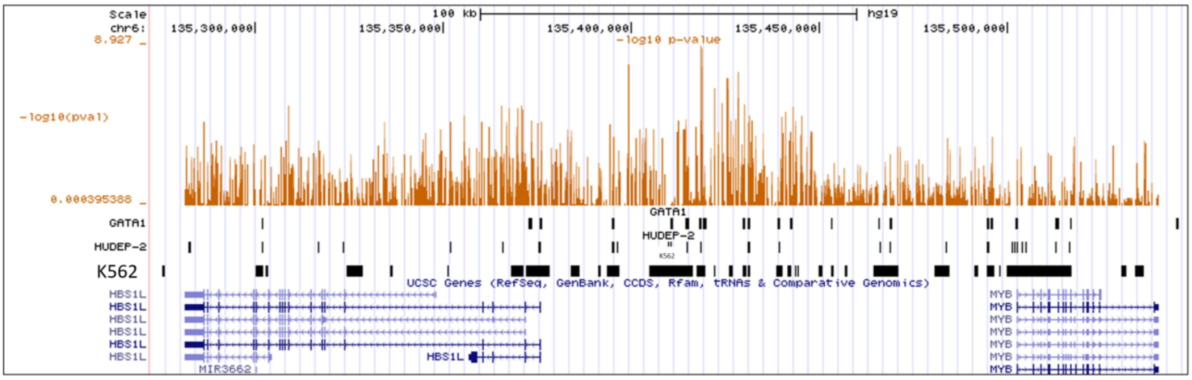


**Supplementary Figure 3:** UCSC plot for the *HMIP* locus. Tracks in order: log10 p-values for variants; GATA1 ChiP-seq annotation; HUDEP-2 annotation; K562 annotation; genes in this locus.

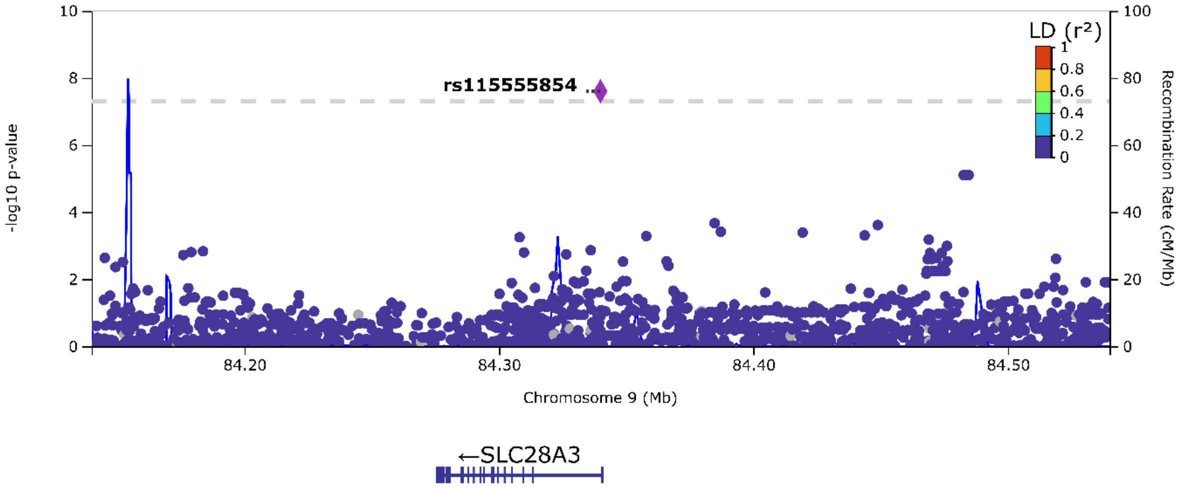


**Supplementary Figure 4.** Regional Association plot of *SLC28A3*

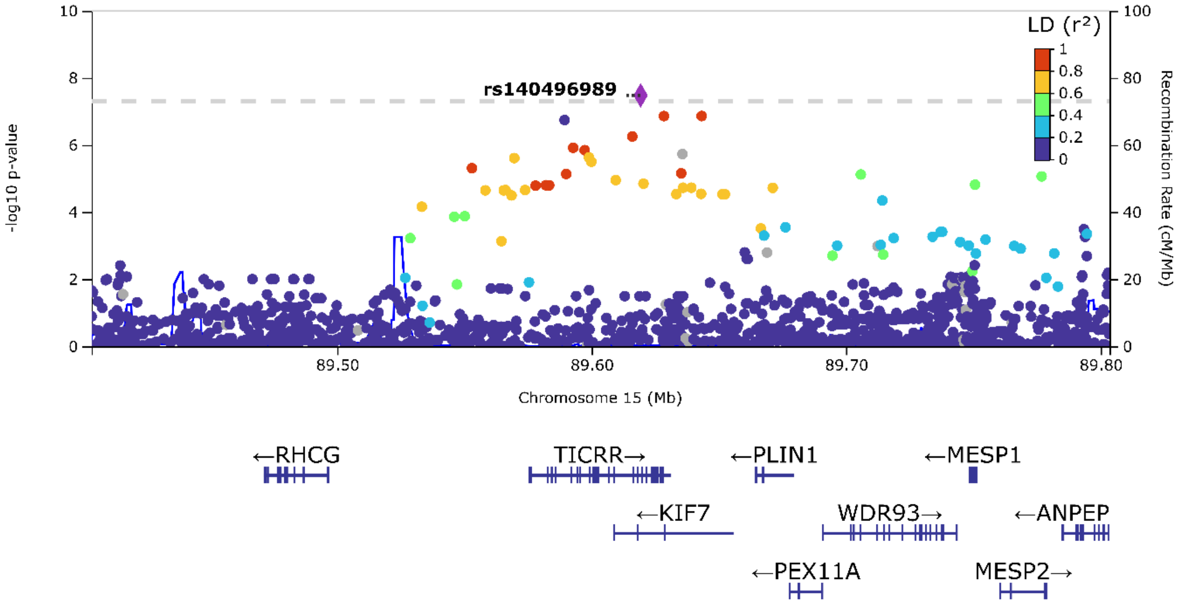


**Supplementary Figure 5.** Regional Association plot of *TICRR*

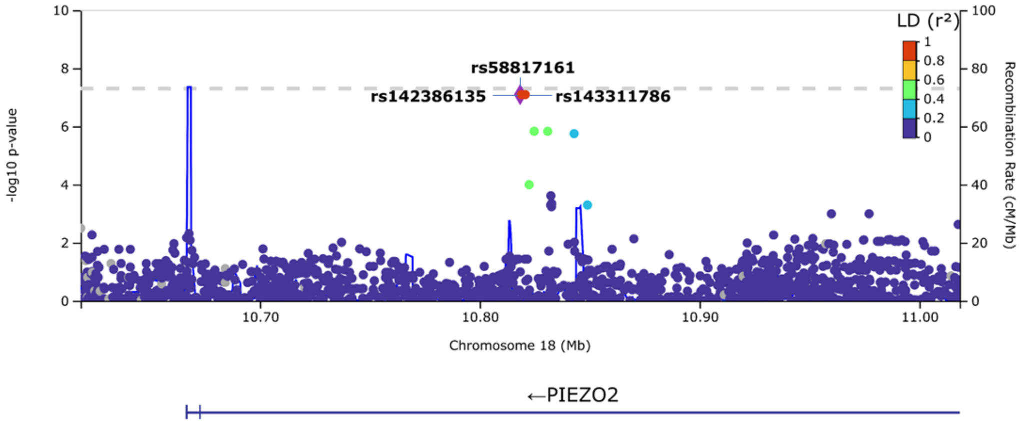


**Supplementary Figure 6.** Regional Association plot of *PIEZO2*

**Supplementary Table 11. Summary statistics of African-American data for replication**

| | | | | | African-American cohort | | | | | | |
|---|---|---|---|---|---|---|---|---|---|---|---|
| | | | | | Cross-sectional (BCM & SCCRIP) | | | Longitudinal (N = 326)[1] | | Longitudinal (N = 326)[2] | |
| **Chr** | **Nearest gene** | **SNP** | **Position** | **EA/OA** | **EAF** | **β** | **P-value** | **β** | **P-value** | **β** | **P-value** |
| 9 | SLC28A3 | rs115555854 | 84339933 | A/C | 0.006 | - | - | -0.53 | 0.16 | -0.38 | 0.25 |
| 11 | SCGB1D4 | rs1406381 | 62301213 | G/T | 0.4482 | -0.05425 | 0.3413 | 0.043 | 0.57 | 0.025 | 0.72 |
| 14 | PSMC6 | rs191465524 | 52721225 | G/T | 0.0147 | -0.1701 | 0.4665 | -0.2 | 0.64 | -0.21 | 0.58 |
| 14 | PSMC6 | rs188152509 | 52740738 | G/C | 0.0147 | -0.1701 | 0.4665 | -0.2 | 0.64 | -0.21 | 0.58 |
| 15 | TICRR | rs78973372 | 89589437 | T/G | 0.0242 | 0.03606 | 0.8364 | 0.27 | 0.19 | 0.23 | 0.22 |
| 15 | TICRR | rs146039045 | 89616003 | A/G | 0.0302 | -0.03462 | 0.8264 | 0.27 | 0.17 | 0.25 | 0.16 |
| 15 | TICRR | rs140496989 | 89619296 | A/G | 0.0242 | 0.03606 | 0.8364 | 0.25 | 0.17 | 0.23 | 0.18 |
| 15 | TICRR | rs137993810 | 89628452 | G/C | 0.0233 | 0.05119 | 0.7731 | 0.31 | 0.14 | 0.27 | 0.16 |
| 15 | TICRR | rs78432130 | 89643261 | G/T | 0.0242 | 0.03606 | 0.8364 | 0.27 | 0.19 | 0.23 | 0.22 |
| 16 | CLCN7 | rs188481235 | 1446101 | T/C | 0.0173 | 0.1487 | 0.4854 | 0.54 | 0.061 | 0.38 | 0.15 |
| 18 | PIEZO2 | rs58817161 | 10818373 | C/T | 0.0164 | -0.06301 | 0.7744 | -2.4 | 0.00071 | -2.5 | 0.00028 |
| 18 | PIEZO2 | rs142386135 | 10819008 | G/A | 0.0138 | -0.1473 | 0.5341 | -2.4 | 0.00071 | -2.5 | 0.00028 |
| 18 | PIEZO2 | rs143311786 | 10820712 | G/A | 0.0164 | -0.06301 | 0.7744 | -0.24 | 0.00071 | -2.5 | 0.00028 |

Chr: Chromosome; SNP: reference ID for the Single Nucleotide Polymorphism; EA: Effect allele; OA: Other allele; EAF: Effect allele frequency; β: allelic effect size; N = sample size; BCM: Baylor College of Medicine cohort; SCCRIP: St. Jude Children' Research Hospital Sickle Cell Clinical Research and Intervention Program.

1: Longitudinal analysis of HbF at ages 1-6: (6 measurements per person, for a total of 1956 data points). The analysis was adjusted for age, sex, Hydroxyurea therapy, principal components 1-5, and the SNP-age interaction; 2: Longitudinal analysis, adjusting for age, sex, Hydroxyurea therapy, principal components 1-5, the SNP-age interaction, and 11-SNP polygenic score for HbF ($PGS^{HbF}$) as detailed in [7].

**Supplementary Table 12. Genome-wide significant and suggestive loci obtained from the meta-analysis.**

| Chr | Locus | SNP | Position | EA/OA | EAF | N | p-value | Direction of effect allele |
|---|---|---|---|---|---|---|---|---|
| 1 | WDR78 | rs77479749 | 66828256 | C/T | 0.089 | 3554 | $5.74 \times 10^{-7}$ | + |
| 2 | BCL11A | rs1427407 | 60490908 | T/G | 0.237 | 3572 | $1.83 \times 10^{-109}$ | + |
| 2 | TEX51 | rs116591810 | 126849099 | T/C | 0.035 | 3546 | $5.54 \times 10^{-7}$ | - |
| 4 | GRID2 | rs539530558 | 93555532 | T/G | 0.01 | 1759 | $4.67 \times 10^{-7}$ | - |
| 5 | AC01460.1 | rs1560177 | 23322878 | C/T | 0.189 | 3562 | $6.5 \times 10^{-7}$ | + |
| 6 | HBS1L-MYB | rs9399137 | 135097880 | C/T | 0.044 | 3562 | $1.77 \times 10^{-27}$ | + |
| 6 | NKAIN2 | rs138298800 | 124625064 | A/G | 0.011 | 1179 | $2.69 \times 10^{-7}$ | - |
| 6 | ATP5MGP2 | rs149569206 | 122875198 | A/T | 0.016 | 3504 | $1.16 \times 10^{-7}$ | - |
| 9 | SLC28A3 | rs115555854 | 84339933 | A/C | 0.015 | 1003 | $2.73 \times 10^{-8}$ | - |
| 11 | OR52S1P | rs4531463 | 5078160 | A/G | 0.074 | 1375 | $2.03 \times 10^{-10}$ | + |
| 14 | AC005520.4 | rs115521884 | 73844597 | C/A | 0.079 | 355 | $9.25 \times 10^{-7}$ | + |
| 17 | FAAP100 | rs12675 | 81540246 | A/G | 0.272 | 2926 | $7.69 \times 10^{-7}$ | + |
| 21 | AF254983.2 | rs79114373 | 10399098 | T/C | 0.117 | 579 | $8.25 \times 10^{-7}$ | - |

Chr: Chromosome; SNP: reference ID for the Single Nucleotide Polymorphism; EA: Effect allele; OA: Other allele; EAF: Effect allele frequency; N = sample size.
